# Neurological and psychiatric presentations associated with human monkeypox virus infection: a systematic review and meta-analysis

**DOI:** 10.1101/2022.07.03.22277069

**Authors:** James B Badenoch, Isabella Conti, Emma R Rengasamy, Cameron J Watson, Matthew Butler, Zain Hussain, Alasdair G Rooney, Michael S Zandi, Glyn Lewis, Anthony S David, Catherine F Houlihan, Ava Easton, Benedict D Michael, Krutika Kuppalli, Timothy R Nicholson, Thomas A Pollak, Jonathan P Rogers

## Abstract

**Objectives:** Neuropsychiatric presentations of monkeypox (MPX) infection have not been well characterised, Despite evidence of nervous system involvement associated with two related *Orthopoxviruses*, in the case of smallpox infection (with the variola virus) and smallpox vaccination (which contains live vaccinia virus). In this systematic review and meta-analysis, we aim to determine the prevalence and describe the spectrum of neurological and psychiatric presentations of MPX.

**Design:** Systematic review and meta-analysis

**Data sources:** MEDLINE, EMBASE, PsycINFO, AMED and pre-print server (MedRxiv) searched up to 31/05/2022

**Eligibility criteria for study selection and analysis:** Any study design of humans infected with MPX that reported neurological or psychiatric presentation. Studies which included more than ten individuals, and symptoms that were reported in a minimum of two separate studies were eligible for meta-analysis

**Data synthesis:** Results were pooled with random-effects meta-analysis to calculate generalised linear mixed models and corresponding 95% confidence intervals for each prevalence outcome.

Heterogeneity was measured with the I^2^ statistic. All included studies are summarised through a narrative synthesis. Risk of bias was assessed with the Newcastle Ottawa Scale and the Joanna Briggs Institute quality assessment tool.

**Results:** From 1,702 unique studies, we extracted data on 19 eligible studies (1,512 participants, 1,031 with confirmed infection using CDC criteria or PCR testing) most of which were cohort studies and case series with no controlled populations. Study quality was generally moderate. Six clinical features were eligible for meta-analysis, of which the most prevalent were myalgia in 55.5% [95%CI 12.1-91.9%], headache 53.8% [30.6-75.4%], fatigue 36.2% [2.0-94.0%], seizure 2.7% [0.6-10.2%], confusion 2.4% [1.1-5.2%] and encephalitis 2.0% [0.5-8.2%]. Heterogeneity significantly varied across clinical features (I^2^=0%-98.7%). Other reported presentations not eligible for meta-analysis included sensory-perceptual disturbance (altered vision, dizziness, and photophobia) and psychiatric symptoms (anxiety and depression).

**Conclusions:** There is preliminary evidence for a range of neurological and psychiatric presentations of MPX, ranging from commonly reported and nonspecific neurological symptoms (myalgia and headache) to rarer but more severe neurological complications, such as encephalitis and seizures. There is less evidence regarding the psychiatric sequelae of MPX, and although there are multiple reports of anxiety and depression the prevalence of these symptoms is unknown. MPX-related nervous system presentations may warrant surveillance within the current MPX outbreak, with prospective longitudinal studies evaluating the mid to long-term sequelae of the virus. Robust methods to evaluate the potential causality of MPX with these clinical features are required at an individual and epidemiological level.

**Systematic review registration:** PROSPERO ID 336649

**SUMMARY BOX:** *What is already known on this topic:*

- Neuropsychiatric symptoms can be highly disabling and have a detrimental effect on quality of life.
- Neuropsychiatric manifestations of monkeypox virus infection have not been well characterised, however, there is evidence of nervous system involvement with the related smallpox virus and vaccinia vaccine.

*What this study adds:*

- Preliminary evidence for a range of neurological and psychiatric presentations of monkeypox infection, ranging from commonly reported and nonspecific neurological symptoms (myalgia and headache) to rarer but more severe neurological complications, such as encephalitis and seizures.
- There is less evidence regarding the psychiatric sequelae of monkeypox infection, and although there are multiple reports of anxiety and depression the prevalence of these symptoms is unknown.
- This preliminary suspicion that there are monkeypox-related nervous system manifestations may warrant both surveillance within the current monkeypox outbreak and robust methods to evaluate the potential causality.

## Introduction

Monkeypox (MPX) is a viral zoonotic disease that belongs to the *Orthopoxvirus* genus of the *Poxviridae* family. MPX was first identified in 1958 in monkeys and rodents in a Danish lab, and human cases were first identified in the Democratic Republic of Congo in 1970^1, 2^. MPX virus has historically been classified in two distinct genetic clades. The Central African (or Congo Basin) clade has been described to be more virulent with a case fatality ratio (CFR) ranging from 1-10% and the West African (WA) clade, less so, with a mortality of < 3%. The WA clade has been identified as the causal agent of the current outbreak^3^. Sporadic outbreaks have occurred outside of its ecological niche, including in the USA in 2003 and the UK in 2018^4, 5^. Since May 13^th^ 2022 a sharp increase in cases, predominantly in the USA and Europe, has brought widened attention to this neglected infectious disease. Concern has arisen due to a high rate of human-to-human transmission and there are current efforts to understand what is driving this transmission^6^. It is unclear what is driving the rising incidence of MPX; however, negligible global levels of immunity to the smallpox virus and its vaccine is a potential factor because smallpox immunity may provide protection against MPX infection^7^.

While dermatological manifestations in the form of a synchronous skin rash in patients with MPX are well documented and characterised, other sequelae such as possible neuropsychiatric effects MPX have yet to be systematically synthesised. Analogous data from smallpox infection and vaccination with Vaccinia (a related *Orthopoxvirus*) indicate that neurological and psychiatric features may be significant. Encephalopathy is a common feature of the clinical presentation of smallpox^8^ and, whilst rare, cases of encephalitis, seizures and stroke have been described following both smallpox infection and vaccination^9, 10^. Encephalitis is estimated to occur in 1 in 500 patients infected with the *Variola major* strain of smallpox and in 1 in 2,000 patients infected with the *Variola minor* strain, occurring 6-10 days after infection^9^. Post-vaccination encephalitis is estimated to occur at a rate of between 2 and 1,219 cases per 100,000 vaccines^11^ with higher rates thought to be associated with use of more neurotropic vaccinia strains^10^, providing *prima facie* support for the relevance of *Orthopoxvirus* biology in the aetiopathology of these sequelae.

In this systematic review and meta-analysis we aimed to (1) summarise the prevalences of neurological and psychiatric presentations of human MPX infection and (2) describe the spectrum of such presentations.

## Methods

This systematic review and meta-analysis was pre-registered on PROSPERO (https://www.crd.york.ac.uk/PROSPERO/display_record.php?RecordID=336649). It is reported according to PRISMA guidelines (checklist is included in Supplementary Table 1).

### Eligibility criteria

Included study types were clinical trials, cohort studies, case-control studies, cross-sectional studies, case series or case reports. Due to the rapidly evolving nature of the literature, pre-prints were included. Studies had to report the prevalence of at least one neurological or psychiatric clinical feature. There were no exclusion criteria based on language. Included studies reported human participants of any age diagnosed with an MPX infection. In order to address our first question about prevalence, studies had to have a minimum of 10 subjects.

### Searches

Ovid was used to search MEDLINE, EMBASE, PsycINFO and AMED without filters or limits up to 31/05/2022. The overall search strategy was to combine terms indicating MPX infection AND terms indicating neurological or psychiatric presentations. Text searches and subject headings were used. The full search strategy is presented in Supplementary Methods 1. MedRxiv was searched for pre-prints published in the previous 12 months. There was manual searching of the reference lists of included papers and other relevant systematic reviews to identify additional relevant studies. Authors in the field were contacted in an attempt to identify unpublished data.

Screening of titles and abstracts for each article was conducted independently by three of the authors (JB, IC, CJW) using Rayyan QCRI (http://www.rayyan.ai/). Where there was disagreement, articles were included for reviewing in the next stage. The list of potentially eligible full texts was imported to a spreadsheet, where two authors (JB, IC) independently assessed eligibility by comparing studies against the eligibility criteria. Where there was disagreement on the inclusion of a full text, a third author (JPR) arbitrated.

### Data extraction

Two of the authors (JB, IC) independently extracted data from each study. Where relevant data were unclear or missing, study investigators were contacted by email for clarification. Where there were discrepancies between reviewers, the two reviewers discussed and agreed on a consensus.

Outcomes were defined as any neurological or psychiatric presentations in individuals infected with human MPX. Data were sought at the level of summary estimates. The specific neurological and psychiatric presentations on which data were collected were derived *post hoc* from the data available in the included papers. All results that were compatible with an outcome in each study were included. Data were also collected for the following study characteristics: study metadata (title, author, citation), country of study population, data collection period, study population, single- vs multicentre, study design, inclusion criteria, exclusion criteria, number with a suspected MPX infection, number in whom MPX infection was confirmed, method of MPX confirmation, number of cases not hospitalised, number of cases hospitalised, number of cases hospitalised and admitted to intensive care, number of cases female, age (mean, SD, median and IQR) of the cases, ethnicity of cases, whether there was a control group, number in the control group, control group description, control group matching parameters, method of identification of neurological or psychiatric presentations, temporality of neurological or psychiatric presentations, number with each available neurological or psychiatric presentation, investigation results, qualitative data, outcome and mortality.

Where an outcome was mentioned in at least one participant in a study, it was assumed that it was not present in any participants in whom it was not mentioned. Where relevant data were only available in graphical representations, e.g., Reynolds et al., manual graphical methods were used to estimate prevalence figures^12^.

### Outcomes, summary measures, and synthesis of results

Results for each outcome were grouped together for analysis. The effect measures sought were period prevalences over the course of the illness. Studies were tabulated in two ways. In one table, each included study was presented sequentially, summarising its design, participants and outcomes. In a second table, results were presented grouped by neurological or psychiatric presentations.

### Meta-analyses

For the meta-analysis, every neuropsychiatric presentation reported by two or more studies was examined. In certain instances there was evidence of overlapping populations between studies, potentially affecting prevalence estimates. To manage this, where overlap was suspected (e.g., Nigeria:^13–16^; USA;^4, 17, 18^ the study with the largest population was included in meta-analysis. However, if for a given presentation (e.g., myalgia or encephalitis) the study with the largest population did not report data for that symptom, the study with the next largest population was chosen for that particular symptom.

Results were pooled with random-effects meta-analysis, using the *metafor* package^19^ in *R* version 4.0.2 to calculate generalised linear mixed models for each prevalence outcome^20, 21^ before using the inverse variance method with the Freeman–Tukey double arcsine transformation as a comparative sensitivity analysis^22^. Between-study heterogeneity was assessed using the *I*² statistic. For interpretation, forest plots were produced with 95% confidence intervals (CIs). Subgroup analyses were planned to investigate heterogeneity where there were five or more included studies for any particular outcome by the following groups: study design (prospective vs retrospective), illness severity, method of diagnosis (serological vs clinical). The threshold for statistical significance was set to p-values of less than 0.05.

### Risk of bias

Risk of bias was assessed using the Newcastle-Ottawa Scale for cohort studies, case-control studies and cross-sectional studies^23^. For case reports and case series, the Joanna Briggs Institute quality assessment tool was used^24^. Two authors assessed each study independently (JB, IC). Where there were discrepancies, a third author arbitrated (ER). Results for each study were presented and patterns in scores analysed. The overall certainty of the evidence was determined by a consideration of the heterogeneity and the risk of bias for an outcome.

### Patient and public involvement

Due to the urgency of this review, patients and members of the public were not involved in the design of this study. The Encephalitis Society, the world’s largest brain inflammation charity, were consulted during the analysis and writing-up stage for assistance in interpretation of the results, and is reflected by Dr Easton’s co-author status.

## Results

The search strategy yielded 2,285 studies. After automatic and manual de-duplication, the titles and abstracts of 1,702 studies were screened and the full texts of 85 studies were assessed for eligibility. An additional seven studies were included from screening references of eligible studies and other relevant systematic reviews. A total of 19 eligible studies were included (Figure 1 - Prisma). Brief reasons for excluding studies are listed in Table S1. Authors were contacted for unpublished data.

**Figure 1:**
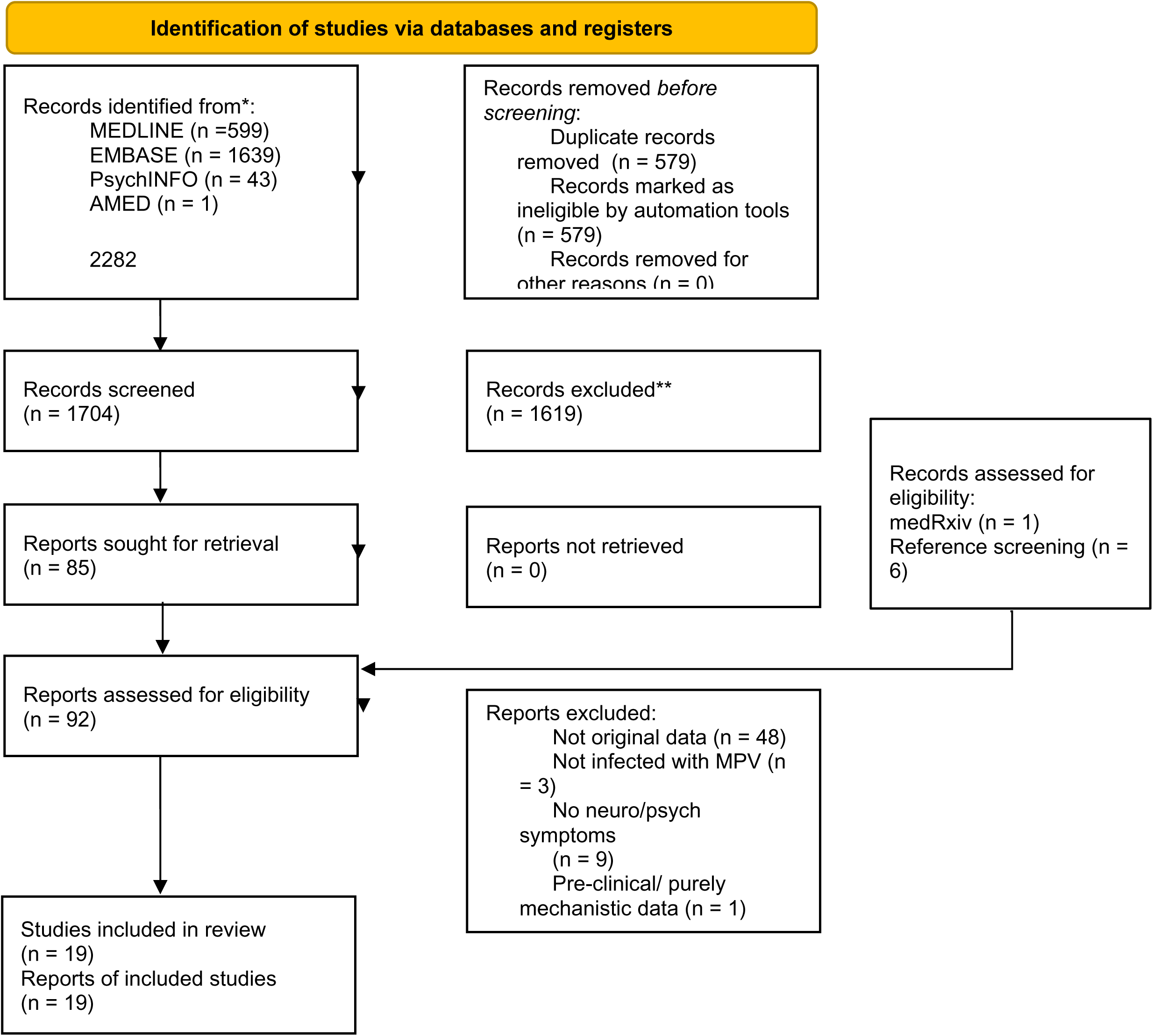
PRISMA flow diagram *Consider, if feasible to do so, reporting the number of records identified from each database or register searched (rather than the total number across all databases/registers). **If automation tools were used, indicate how many records were excluded by a human and how many were excluded by automation tools. *From:* Page MJ, McKenzie JE, Bossuyt PM, Boutron I, Hoffmann TC, Mulrow CD, et al. The PRISMA 2020 statement: an updated guideline for reporting systematic reviews. BMJ 2021;372:n71. doi: 10.1136/bmj.n71

### Population and study characteristics

The 19 studies included a total population *n*=1,512 individuals (sample size range 1-370; median *n*=21) with suspected MPX, *n*=1,031 (68.9%) of whom had infection confirmed by PCR. The mean (SD) age reported was 24.219.2) years, based on only 8 studies (n=542). Just under half of the population was female (n=465, 44.8%). Among studies reporting the setting of MPX treatment (12 studies, n=390), most patients were hospitalised (n=331, 84.9%). Ethnicity was explicitly reported in only three studies (n=54) in whom all were from the USA and 46 (85%) were White. Most studies originated in the USA (six studies) followed by Nigeria, the Democratic Republic of Congo (five studies each), the Republic of Congo (two studies), and the UK (one study) (Table 1).

**Table 1:** Characteristics of included subjects and studies

|  |  |
| --- | --- |
| **Characteristics** |  |
| Sample size (range, median) | 1-370 (21) |
| Confirmed Monkeypox (PCR) (n, %) | 1031/1512 (68.2) |
| Age (mean, SD) | 24.2 (19.4) |
| Sex: female (n, %) | 465/1039 (44.80%) |
| **Location of MPV treatment** (n, %) | 331/390 (84.9) |
| Hospital | 88/390 (22.6) |
| Community | 641 |
| Not stated |  |
| **Country** (n) |  |
| USA | 6 |
| Democratic Republic of Congo | 5 |
| Republic of Congo | 2 |
| UK | 1 |
| **Ethnicity** (n) |  |
| Stated | 54 |
| Unstated | 977 |
| White | 46 |
| Asian |  |
| Black African |  |
| Black non-African | 1 |
| Mixed / multiple |  |
| Hispanic | 1 |
| Other (Arab or other) |  |
| **Study Design** (n) |  |
| Cohort | 12 |
| Cross-sectional | 2 |
| Case series | 4 |
| Case report | 1 |
| Retrospective* | 7 |
| Prospective* | 7 |
| Single centre | 11 |
| Multicentre | 7 |
| Unclear | 1 |
| **NOS Quality Assessment\*\*** |  |
| Low | 6 |
| Medium | 8 |
| High | 0 |

\*only applies to cohort and cross-sectional studies
\*\*Based on 14 studies (NOS used for cohort and cross-sectional studies only)

Most studies (12/19) had a cohort design, two were cross-sectional and the remainder were case series (four) and one case report. Only one study included a comparison group^25^. There was an equal split of prospective and retrospective cohort and cross-sectional studies.

Study quality scores were assessed using the Newcastle-Ottawa Scale (cohort and cross-sectional studies) and Joanna Briggs Quality Assessment Tool (case series and reports) are summarised in Table 2. Regarding the former, studies were generally scored down for no points on comparability due to a lack of control group in all bar one of the included studies. Furthermore, a lack of reported follow-up for the majority of studies also reduced the outcome score on the Newcastle Ottawa.

**Table 2:** Quality Assessment Scores

A: Quality Assessment with Newcastle-Ottawa Scale (Cohort and cross-sectional studies)
| Study | Design | Selection (4) | Comparability (3) | Outcome (3) | Total |
| --- | --- | --- | --- | --- | --- |
| Ogoina et al., 202013 | Cohort | *** | - | ** | 5 |
| Huhn et al.,200525 | Cohort | *** | - | * | 4 |
| Yinka-Ogunleye et al. 201915 | Cohort | **** | - | * | 5 |
| Boumandouki et al., 200729 | Cohort | * | - | * | 2 |
| Akar et al., 202016 | Cohort | * | - | * | 2 |
| Croft et al., 200728 | Cohort | ** | - | *** | 5 |
| Pittman et al., 202232 | Cohort | ** | - | ** | 4 |
| Adler et al., 20225 | Cohort | ** | - | * | 3 |
| Reed et al., 200417 | Cohort | ** | - | * | 3 |
| Reynolds., 200618 | Cohort | ** | - | * | 3 |
| Ježek et al., 198730 | Cohort | *** | - | ** | 5 |
| Kalthan et al., 201633 | Cohort | ** | - | * | 3 |
| Ogoina et al., 201914 | Cross-sectional | **** | - | ** | 6 |
| Hughes et | Cross- | *** | - | *** | 6 |
| al., 2021<sup>25</sup> | sectional |  |  |  |  |

Number of asterisks indicates total score for a domain out of the bracketed total in the column heading. B: Quality Assessment with Joanna Briggs Quality Assessment Tool (case series)
| Study | Inclusion criteria<sup>a</sup> | Measurement of condition<sup>b</sup> | Identification of condition<sup>c</sup> | Consecutive inclusions<sup>d</sup> | Complete inclusion of participants<sup>e</sup> | Reporting of participant demographics<sup>f</sup> | Reporting of clinical information<sup>g</sup> | Outcome reporting<sup>h</sup> | Presenting site(s)/clinic(s) demographics<sup>i</sup> | Statistical analysis appropriate<sup>j</sup> |
| --- | --- | --- | --- | --- | --- | --- | --- | --- | --- | --- |
| Learned et al., 2005<sup>34</sup> | * | * | * | * | * | * | * | * | * | - |
| Ježek et al., 1987<sup>30</sup> | - | * | * | - | - | - | - | - | * | - |
| Sejvar et al., 2004<sup>26</sup> | * | * | * | - | * | * | * | * | * | - |
| Reynolds et al., 2006<sup>18</sup> | * | * | - | * | * | * | * | - | * | - |

\* indicates a domain was met. No studies had statistical analysis, so the domain was not relevant.
<sup>a</sup>Were there clear criteria for inclusion in the case series?
<sup>b</sup>Was the condition measured in a standard, reliable way for all participants included in the case series?
<sup>c</sup>Were valid methods used for identification of the condition for all participants included in the case series?
<sup>d</sup>Did the case series have consecutive inclusion of participants?

| **Domain** | **Outcome** |
| --- | --- |
| Were patient's demographic characteristics clearly described? | Yes |
| Was the patient's history clearly described and presented as a timeline? | Yes |
| Was the current clinical condition of the patient on presentation clearly described? | Yes |
| Were diagnostic tests or assessment methods and the results clearly described? | Yes |
| Was the intervention(s) or treatment procedure(s) clearly described? | Yes |
| Was the post-intervention clinical condition clearly described? | No |
| Were adverse events (harms) or unanticipated events identified and described? | Yes |
| Does the case report provide takeaway lessons? | Yes |

Study populations were mostly drawn from national case surveillance projects (e.g., Nigeria:^15, 16^.; USA:^4, 17^ or cohort studies evaluating the same outbreak of MPX^18, 26, 27^ Table 3. Other populations were more selective, including a sample of individuals co-infected with Varicella zoster virus^25^ or an evaluation of veterinary workers exposed to an infected prairie dog[28]. All studies confirmed MPX infection with PCR, except for Boumandouki et al^29^, and Centre for Disease Control (CDC) definitions of confirmed cases were followed in most studies. Nine studies reported the clade of MPX isolated in infected individuals. Of these, the majority were West African variants including all six studies in the USA. Two studies reported smallpox vaccination status^29, 30^ of which, the latter found deaths from MPX infection were confined to those not vaccinated for smallpox.

**Table 3.**
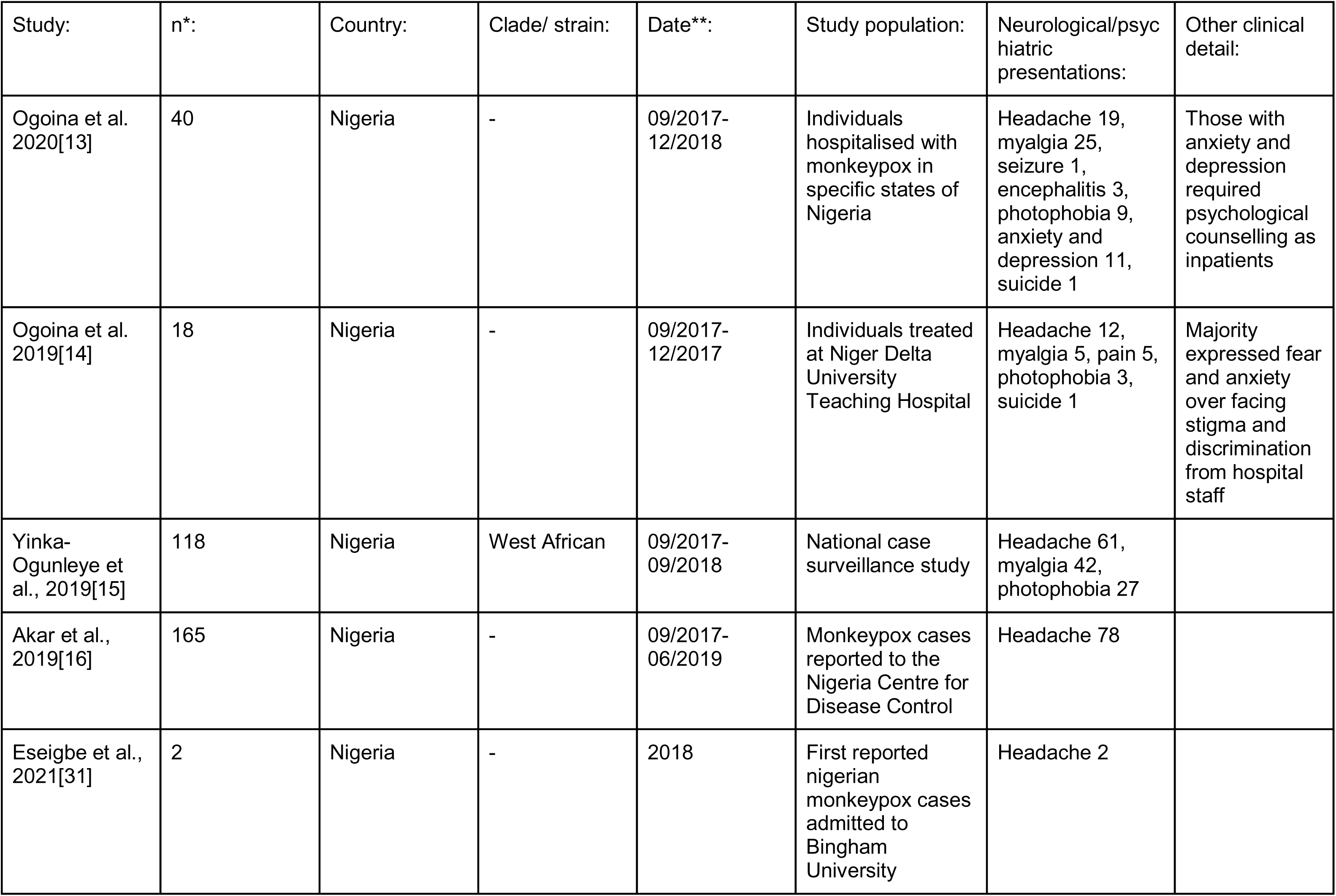
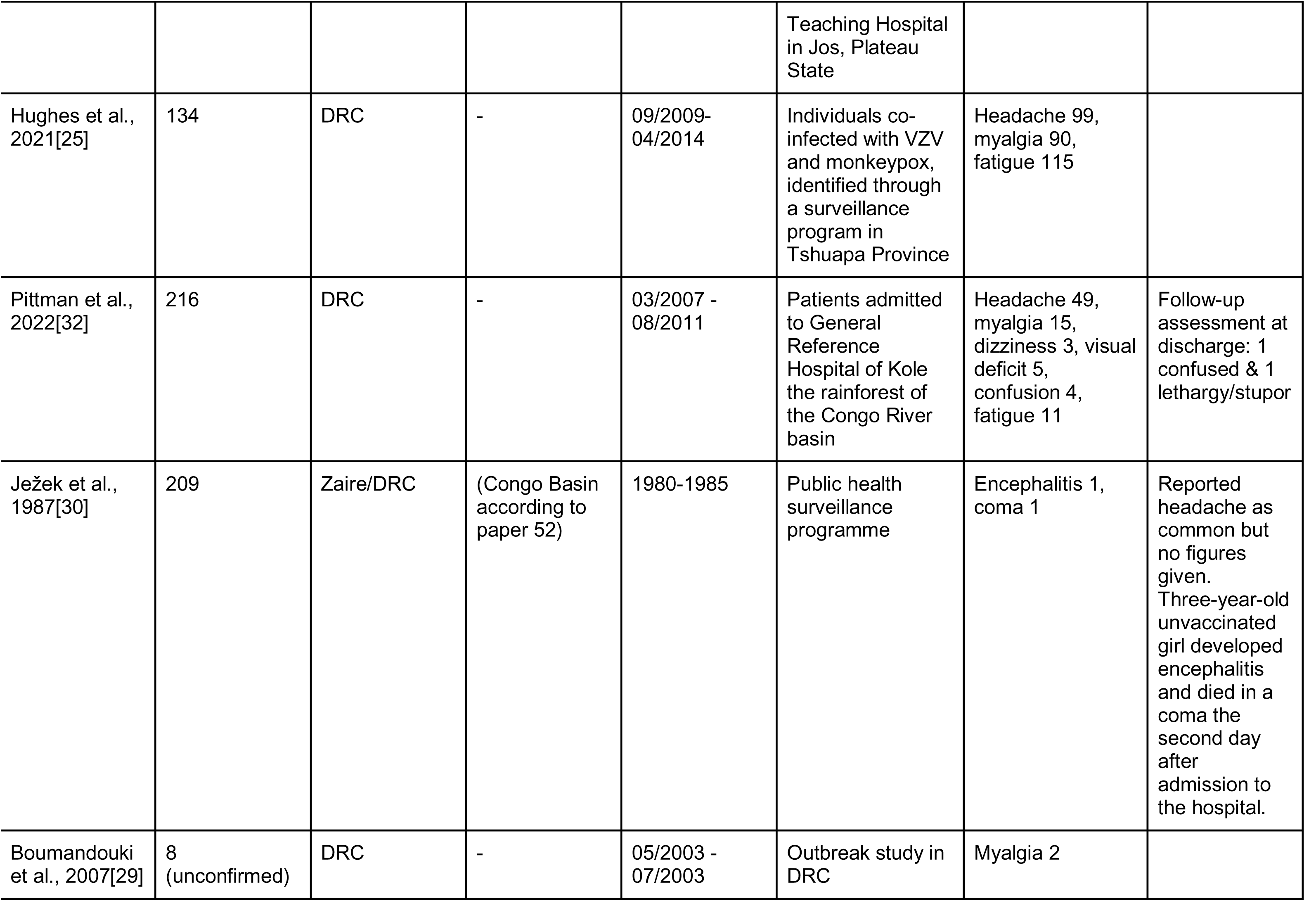
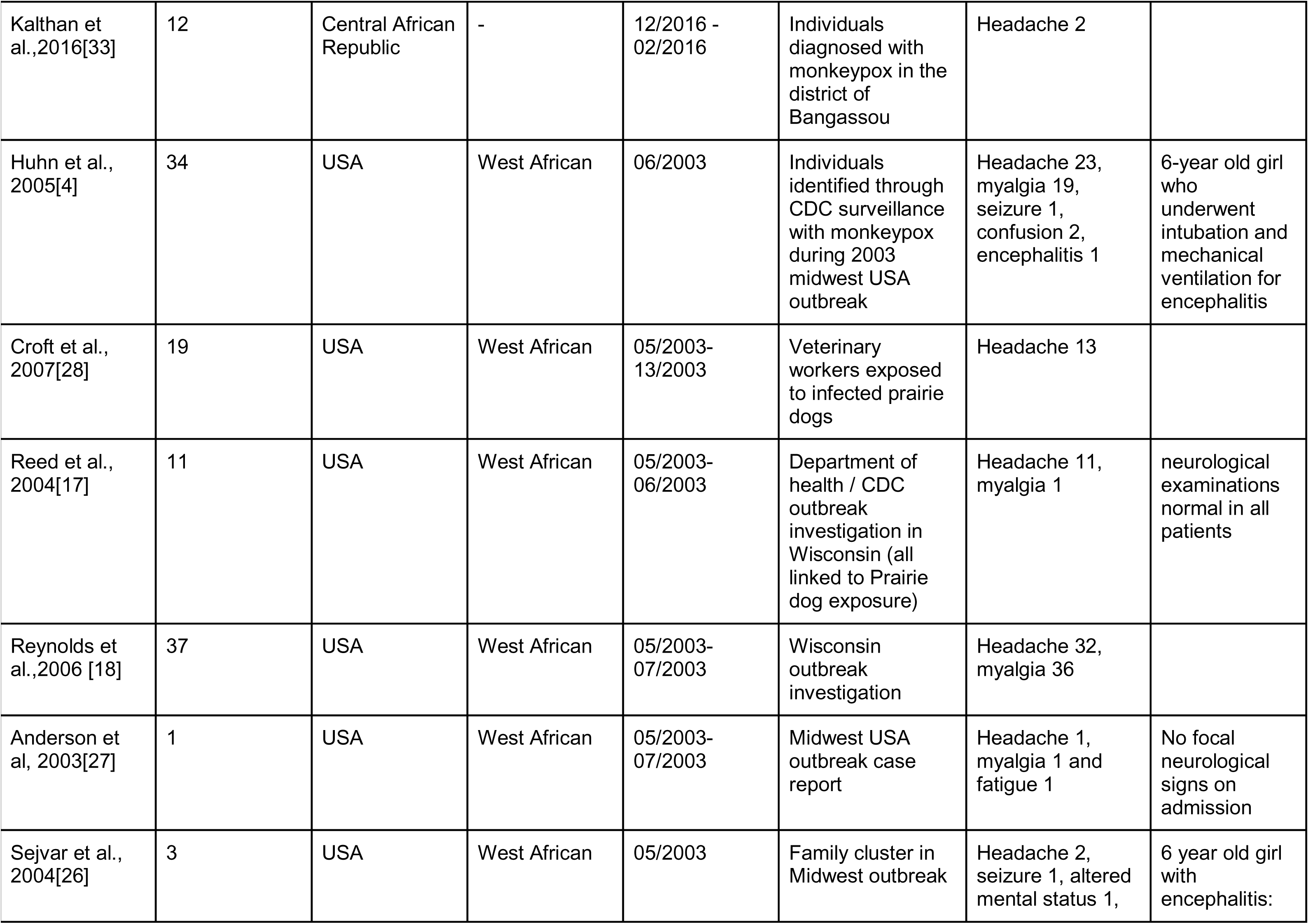
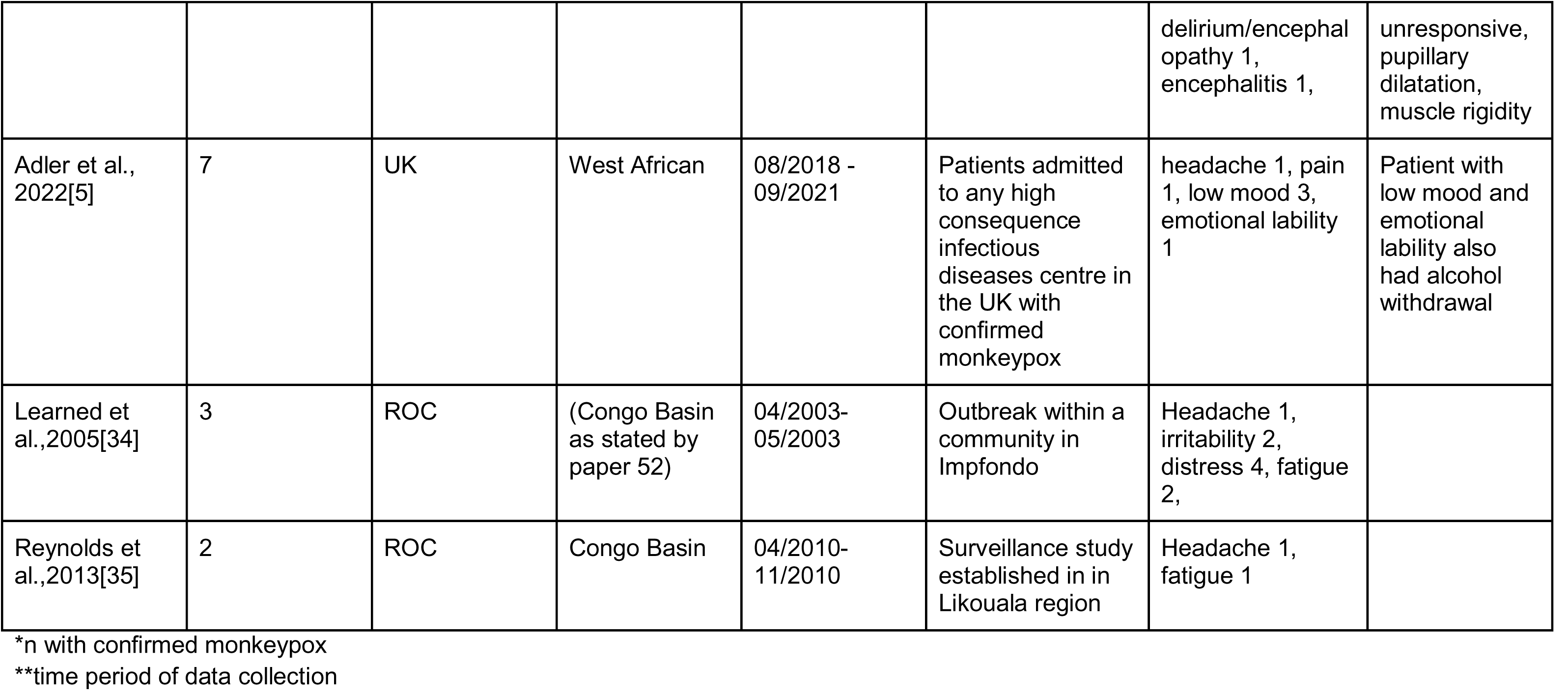
summary of included studies

| Study: | n*: | Country: | Clade/ strain: | Date**: | Study population: | Neurological/psychiatric presentations: | Other clinical detail: |
| --- | --- | --- | --- | --- | --- | --- | --- |
| Ogoina et al. 2020[13] | 40 | Nigeria | - | 09/2017-12/2018 | Individuals hospitalised with monkeypox in specific states of Nigeria | Headache 19, myalgia 25, seizure 1, encephalitis 3, photophobia 9, anxiety and depression 11, suicide 1 | Those with anxiety and depression required psychological counselling as inpatients |
| Ogoina et al. 2019[14] | 18 | Nigeria | - | 09/2017-12/2017 | Individuals treated at Niger Delta University Teaching Hospital | Headache 12, myalgia 5, pain 5, photophobia 3, suicide 1 | Majority expressed fear and anxiety over facing stigma and discrimination from hospital staff |
| Yinka-Ogunleye et al., 2019[15] | 118 | Nigeria | West African | 09/2017-09/2018 | National case surveillance study | Headache 61, myalgia 42, photophobia 27 |  |
| Akar et al., 2019[16] | 165 | Nigeria | - | 09/2017-06/2019 | Monkeypox cases reported to the Nigeria Centre for Disease Control | Headache 78 |  |
| Eseigbe et al., 2021[31] | 2 | Nigeria | - | 2018 | First reported nigerian monkeypox cases admitted to Bingham University | Headache 2 |  |
|  |  |  |  |  | Teaching Hospital in Jos, Plateau State |  |  |
| Hughes et al., 2021[25] | 134 | DRC | - | 09/2009-04/2014 | Individuals co-infected with VZV and monkeypox, identified through a surveillance program in Tshuapa Province | Headache 99, myalgia 90, fatigue 115 |  |
| Pittman et al., 2022[32] | 216 | DRC | - | 03/2007 - 08/2011 | Patients admitted to General Reference Hospital of Kole the rainforest of the Congo River basin | Headache 49, myalgia 15, dizziness 3, visual deficit 5, confusion 4, fatigue 11 | Follow-up assessment at discharge: 1 confused & 1 lethargy/stupor |
| Ježek et al., 1987[30] | 209 | Zaire/DRC | (Congo Basin according to paper 52) | 1980-1985 | Public health surveillance programme | Encephalitis 1, coma 1 | Reported headache as common but no figures given. Three-year-old unvaccinated girl developed encephalitis and died in a coma the second day after admission to the hospital. |
| Boumandouki et al., 2007[29] | 8 (unconfirmed) | DRC | - | 05/2003 - 07/2003 | Outbreak study in DRC | Myalgia 2 |  |
| Kalthan et al.,2016[33] | 12 | Central African Republic | - | 12/2016 - 02/2016 | Individuals diagnosed with monkeypox in the district of Bangassou | Headache 2 |  |
| Huhn et al., 2005[4] | 34 | USA | West African | 06/2003 | Individuals identified through CDC surveillance with monkeypox during 2003 midwest USA outbreak | Headache 23, myalgia 19, seizure 1, confusion 2, encephalitis 1 | 6-year old girl who underwent intubation and mechanical ventilation for encephalitis |
| Croft et al., 2007[28] | 19 | USA | West African | 05/2003-13/2003 | Veterinary workers exposed to infected prairie dogs | Headache 13 |  |
| Reed et al., 2004[17] | 11 | USA | West African | 05/2003-06/2003 | Department of health / CDC outbreak investigation in Wisconsin (all linked to Prairie dog exposure) | Headache 11, myalgia 1 | neurological examinations normal in all patients |
| Reynolds et al.,2006 [18] | 37 | USA | West African | 05/2003-07/2003 | Wisconsin outbreak investigation | Headache 32, myalgia 36 |  |
| Anderson et al, 2003[27] | 1 | USA | West African | 05/2003-07/2003 | Midwest USA outbreak case report | Headache 1, myalgia 1 and fatigue 1 | No focal neurological signs on admission |
| Sejvar et al., 2004[26] | 3 | USA | West African | 05/2003 | Family cluster in Midwest outbreak | Headache 2, seizure 1, altered mental status 1, | 6 year old girl with encephalitis: |
|  |  |  |  |  |  | delirium/encephalopathy 1, encephalitis 1, | unresponsive, pupillary dilatation, muscle rigidity |
| Adler et al., 2022[5] | 7 | UK | West African | 08/2018 - 09/2021 | Patients admitted to any high consequence infectious diseases centre in the UK with confirmed monkeypox | headache 1, pain 1, low mood 3, emotional lability 1 | Patient with low mood and emotional lability also had alcohol withdrawal |
| Learned et al.,2005[34] | 3 | ROC | (Congo Basin as stated by paper 52) | 04/2003-05/2003 | Outbreak within a community in Impfondo | Headache 1, irritability 2, distress 4, fatigue 2, |  |
| Reynolds et al.,2013[35] | 2 | ROC | Congo Basin | 04/2010-11/2010 | Surveillance study established in in Likouala region | Headache 1, fatigue 1 |  |

\*n with confirmed monkeypox
\*\*time period of data collection

Furthermore, mortality was reported in ten studies and varied between 0-25% in studies with 10 or more individuals. Neurological and psychiatric presentations varied widely, however, the most frequently reported were headache, myalgia, seizure, confusion, encephalitis and fatigue (table 3). Neuropsychiatric features were mostly evaluated through case note review in retrospective studies and a mix of clinical interview and questionnaire in prospective studies. The breadth of clinical features assessed in the latter design was minimal. For example, in two prospective studies, the only neuropsychiatric presentations evaluated were headache, fatigue and myalgia^15^; Croft et al^28^ headache only. Assessment of clinical feature severity, using standardised scales, and chronicity was also lacking.

### Prevalence of neurological and psychiatric presentations

After exclusion of potentially overlapping populations, six neuropsychiatric presentations were eligible for meta-analysis of prevalence. The most frequent clinical feature was myalgia (pooled prevalence=55.5% [95%CI 12.1-91.9%]), followed by headache (53.8% [30.6-75.4%]), fatigue (36.2% [2.0-94.0%]), seizure (2.7% [0.6-10.2%]), confusion (2.4% [1.1-5.2%]) and encephalitis (2.0% [0.5-8.2%]) (Figure 2 – forest plots, Table 4 – prevalence of neurological and psychiatric presentations). Heterogeneity significantly varied across clinical features (*I*^2^=0%-98.7%, Table 4). Other neuropsychiatric presentations including dizziness, pain, altered vision, encephalopathy, photophobia, depression, anxiety and suicide are summarised in table 3.

**Table 4.** Pooled prevalence of individual neurological and psychiatric presentations

| Clinical feature | Pooled Prevalence (%) | CI (%) | number of individuals (n) | number of studies (k) | heterogeneity (%) |
| --- | --- | --- | --- | --- | --- |
| Myalgia | 55.5 | 12.1 - 91.9 | 505 | 4 | 98.7 |
| Headache | 53.8 | 30.6 - 75.4 | 583 | 6 | 95.5 |
| Fatigue | 36.2 | 2.0 - 94.0 | 350 | 2 | 98.6 |
| Seizure | 2.7 | 0.6 - 10.2 | 74 | 2 | 0 |
| Confusion | 2.4 | 1.1 - 5.2 | 250 | 2 | 0 |
| Encephalitis | 2 | 0.5 - 8.2 | 283 | 3 | 55.8 |

**Figure 2.**
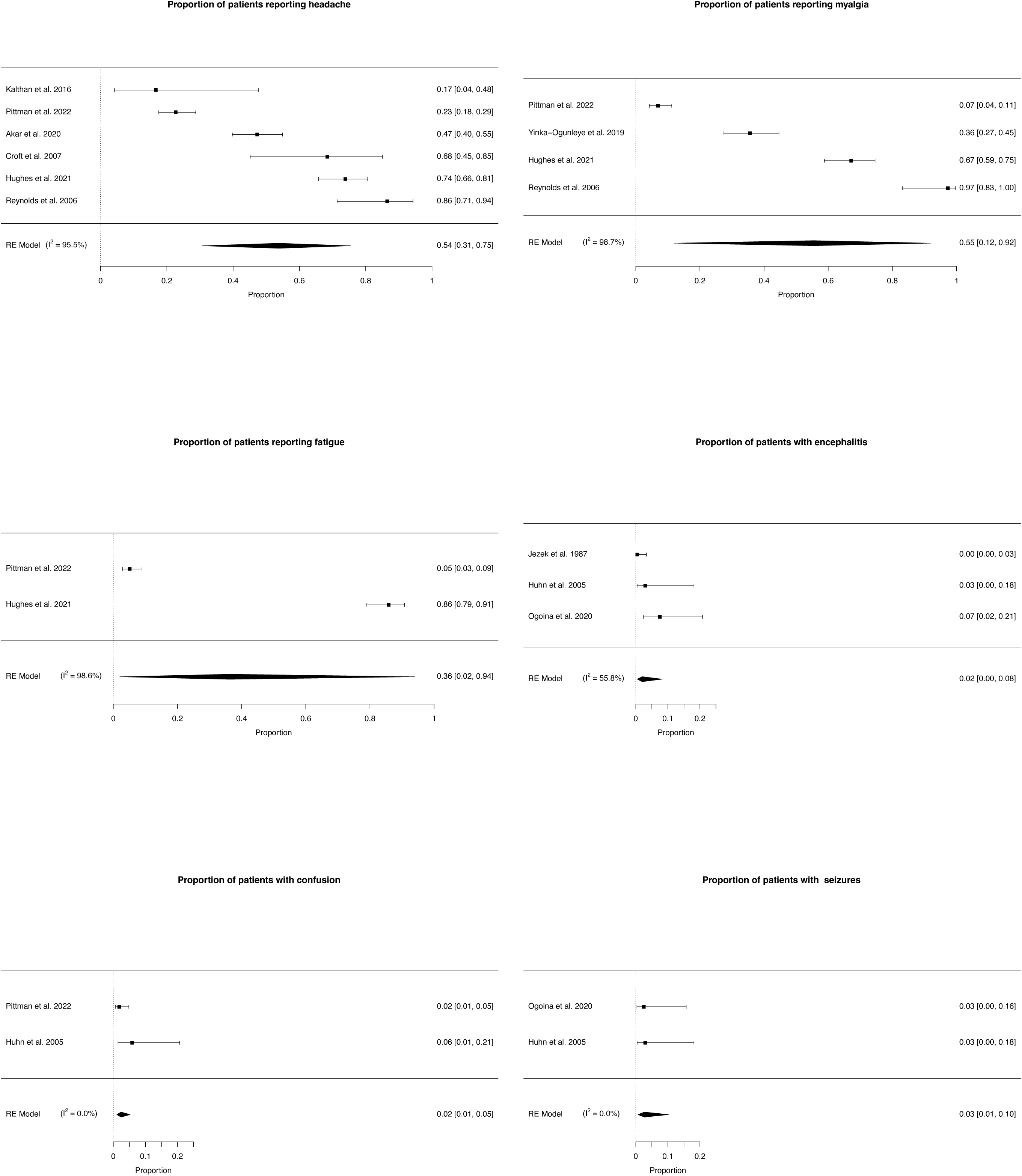
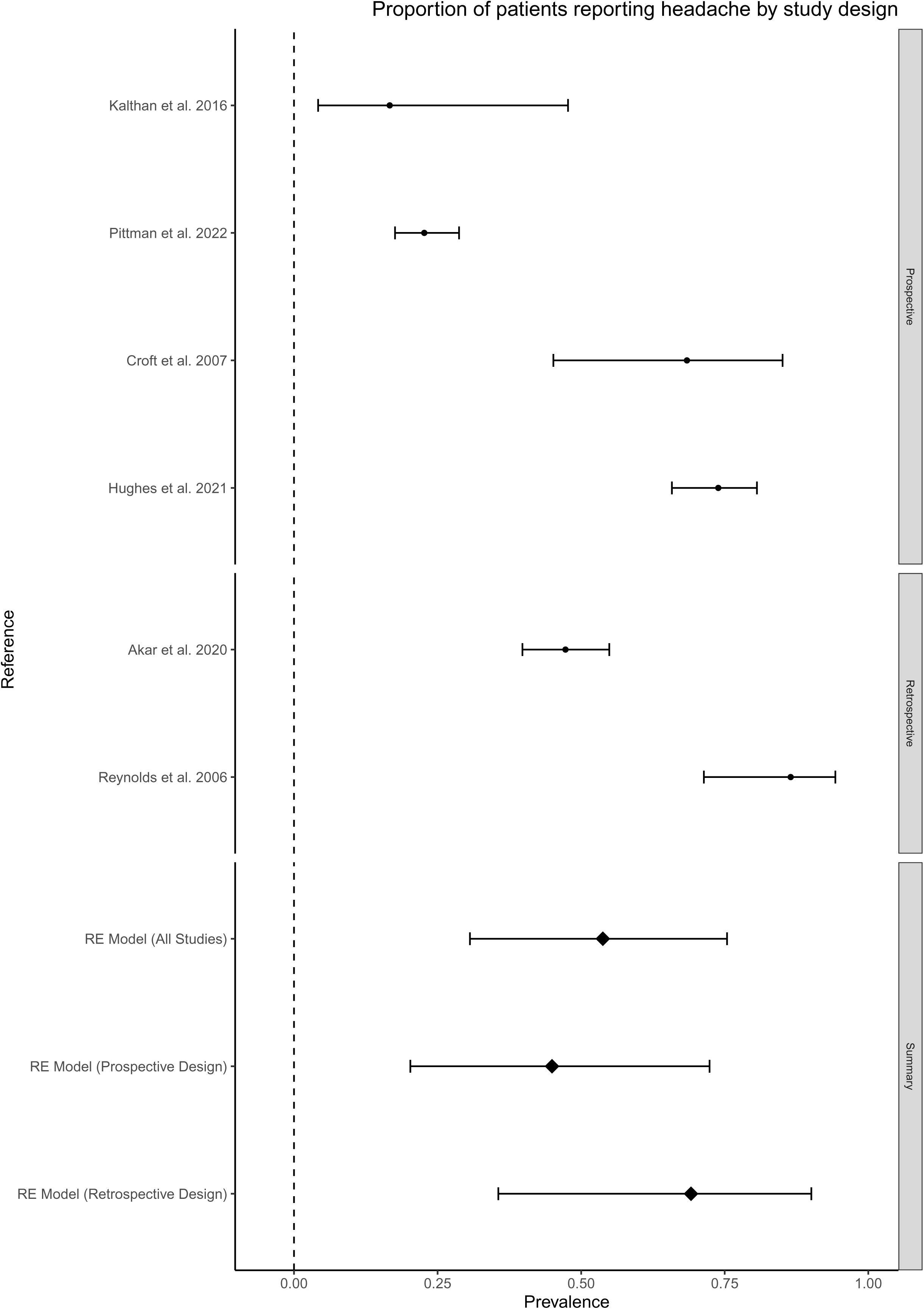
Forest plots for individual neurological and psychiatric manifestations.

### Secondary analysis

There was no statistical evidence for a difference in the prevalence of headache in prospective compared to retrospective studies (based on four and two studies respectively – Figure 3 subgroup analysis of headache). It was not possible to analyse subgroups based on illness severity or method of diagnosis due to missing data and lack of variation between groups. No other clinical features met our prespecified eligibility criteria for subgroup analysis.

## Discussion

This systematic review and meta-analysis provides the first comprehensive overview of the prevalence of neurological and psychiatric presentations of MPX infection. Based on a small number of studies examining this topic, headache and myalgia are present in over half of individuals and more severe central nervous system complications, including encephalitis and seizure, are present in small (<3%) but non-negligible proportions of infected individuals. The prevalence of other neuropsychiatric symptoms including fatigue, anxiety and depression are less clear. There are also knowledge gaps surrounding putative factors which influence risk of neurological and psychiatric presentations including overall MPX illness severity and viral clade.

The relatively high prevalence of non-specific symptoms such as headache and myalgia is perhaps unsurprising given that these symptoms are common in viral infections^36–38^. It is likely that these symptoms represent a reaction to systemic illness rather than direct neurological injury. Additionally, a paucity of follow-up and lack of evaluation of symptom severity and timing makes it hard to ascertain whether these symptoms are potentially highly disabling or milder and/or transient. These findings are consistent with a recently published review of MPX epidemiology which found that fatigue/asthenia and headache were present in over a fifth of individuals and myalgia in slightly fewer^39^. Although less frequently assessed, there was some evidence of psychiatric symptoms in people with MPX. For example, Ogoina and colleagues (2020)^13^ found that psychiatric manifestations including anxiety and depression were present in over a quarter of individuals hospitalised with MPX in Nigeria. Similarly, a case series from specialist centres in the UK found that three of seven patients admitted to hospital suffered from low mood^5^. Although severity data was not reported, in both studies individuals with psychiatric symptoms required inpatient psychological therapies. It is important to note that anxiety and depression are common in hospitalised patients, and indeed in those who are in quarantine for infectious diseases, and the majority of individuals in this review were drawn from quarantined hospitalised samples^40, 41^. This evidence could point to an underrecognized and understudied burden of psychiatric complications in the acute phase of MPX.

The quality of evidence included in this review may reflect the relatively understudied phenomena of nervous system presentations in the context of MPX and affects what conclusions can be drawn. Only one study included a control group, where individuals with Varicella zoster virus (VZV) and MPX were compared to those with MPX only and with VZV only^25^. Individuals with MPX and VZV co-infection were more likely to report fatigue than those with VZV alone, however, no comparative data were given for the MPX only group and no other neuropsychiatric symptoms of interest were compared. However, the clinical manifestations of VZV and MPX co-infection are complicated and the differences between individuals with co-infection and those with VZV alone may not be solely attributable to the effect of MPX. Attributing causality of viral infection to neurological symptoms is difficult despite established criteria used to define it^42^. This is exacerbated by a lack of adequate comparison groups, such as healthy controls or individuals with other viral illnesses. Additionally, incidence cannot be ascertained without reporting of premorbid neurological or psychiatric diagnoses. Small sample sizes also reduce the reliability of prevalence estimates.

Half of the included studies were retrospective and relied on case note review, which risks a systematic under-representation of symptoms, especially, if neuropsychiatric features were not routinely inquired about or assessed, although this is likely to be less of an issue with severe neurological complications. Additionally, no studies included in this review assessed psychiatric symptoms using standardised scales. The clinical significance of these symptoms is thus difficult to ascertain. In terms of data synthesis, we were limited by a lack of reporting of certain variables including MPX severity, ethnicity, and clade of MPX. The small number of studies means that subgroup analysis should be considered purely exploratory. Limited reporting of neurological investigations such as CSF analysis and neuroimaging also hinder understanding of the pathogenesis and potential mechanisms underlying the presentations described.

Though there has been little experimental work conducted on MPX and the nervous system in humans, a small number of case reports looking at smallpox have pointed to several diverse mechanistic explanations. Post-mortem examination revealed acute perivenular demyelination in patients known to have died of smallpox^43^. Additionally, MRI scans in those with post-vaccination encephalitis have been suggestive of acute disseminated encephalomyelitis (ADEM)^10^. However, to date CSF from patients with post-vaccine CNS complications has overwhelmingly been normal with no viral load detected, consistent with aseptic meningitis^9^, pointing to an immune-mediated pathogenesis. However, caution is required in extrapolating from either variola or vaccinia effects or neuropathology to MPX, despite shared genetics and clinical overlap between these *Orthopoxviruses* and their respective clinical syndromes. One case report included in this review of a child with MPX encephalitis, could not isolate viral material from CSF but did detect MPX specific IgM antibodies in CSF^17, 26^. This may suggest an intrathecal immune-mediated response; however, other cases of MPX-encephalitis did not report results of CSF analysis^13^. The underlying mechanisms of MPX neuropsychiatric manifestations include a direct CNS infection, an immune mediated response and a psychological reaction to illness.

Stigma could play a role in maladaptive psychological processes in those with MPX. Several studies emphasise the stigma associated with a diagnosis of MPX both on the individual, their family and integration back into society. Low mood was a common feature seen amongst many infected with MPX^13^. One patient died from suicide a few days after admission. The reports cited worries regarding how he had contracted MPX, and the effects on both him and his family^13^. Others highlight the stigma associated with the focus on transmission related to close physical contact and sexual contact, which may place an unwarranted and potentially harmful emphasis on the LGBTQ+ community. Contemporary public health and education should make clear that although there have been a high number of cases in men who have sex with men, and some cases of MPX with co-infection of HIV/AIDS^15^, MPX can also be spread via direct contact, clothing, and respiratory secretions, and that anyone can become infected. Nevertheless Bragazzi et al, 2022^39^ point out the potential for exacerbation of stigma in already stigmatised communities.

Viral infections are known to have profound psychological effects on those affected, such as fear, loss, discrimination and stigma[44]. Though the clinical course varies amongst individuals, a common progression of dermatological change is persistent scarring. Ogoina, 2020^13^ report that not only were skin lesions widespread, itchy and tender causing disfigurement, but that patients may also develop genital ulcers which were particularly distressing. Meta-analysis indicates a significant burden of persistent anxiety and depression in patients with facial scarring^45^. In addition, Rumsey and Harcourt^46^ highlight the wider negative consequences such as reduced self-esteem and loss of identity. Whilst the studies included in this review focus on acute psychological symptoms, the long-term psychological consequences of MPX infection are unknown.

Similarly, it is unclear what the long-term outcomes for patients with MPX encephalitis are, aside from one reported death^30^. Given that encephalitis, of infectious or autoimmune aetiology, results in considerable neurological and neuropsychiatric morbidity^47^, collecting longitudinal data on affected individuals with this rare complication should be a high priority moving forward. The long-term neurocognitive effects of MPX infection also remain elusive. Pittman and colleagues (2022)^32^ reported a case of confusion and lethargy still present at discharge. Given the range of neuropsychiatric effects that occur in a proportion of people after several viral illnesses^48–50^ it may be worthwhile ascertaining whether these symptoms persist in MPX.

This paper has research and therapeutic implications. The variability in detection and reporting of neuropsychiatric manifestations highlights the need for registries of emerging zoonotic infections where clinicians can provide case histories and reliable data in rapidly evolving epidemics such as the WHO clinical data platform^51^. The CoroNerve surveillance study^52^ demonstrates the utility of rapid reporting, having proved successful in the COVID-19 pandemic. Aside from epidemiology, there are therapeutic implications of this review. Our results suggest it would be worth researching the value of integrating psychological support into the care of those isolated with MPX both in the acute setting and beyond, including those managed in the community. The inclusion of encephalitis as well as the psycho-social and emotional impacts for patients of contracting MPX will likely have implications for patient quality of life and therefore increased research in this field is an important area yet to be adequately addressed for patients and their caregivers/families.

There is preliminary evidence for a range of neurological and psychiatric presentations of MPX, ranging from commonly reported and nonspecific neurological symptoms (myalgia and headache) to rarer but more severe neurological complications, such as encephalitis and seizures. There is less evidence regarding the psychiatric sequelae of MPX, and although there are multiple reports of anxiety and depression the prevalence of these symptoms is unknown. This preliminary suspicion that there are MPXV-related nervous system presentations may warrant surveillance within the current MPX outbreak, with prospective longitudinal studies evaluating the mid to long-term sequelae of the virus. well-powered prospective longitudinal studies to evaluate multi-system MPX effects. Robust methods to evaluate the potential causality of MPXV with these manifestations are required at an individual and epidemiological level.

## Data Availability

The code created and used for the meta-analysis is available on GitHub (link included)

https://github.com/CameronWatson2020/monkeypox/blob/main/monkeypox_analysis.R

---

**Supplementary material: PRISMA 2020 Checklist**

Supplementary material: PRISMA 2020 Checklist

| Section and Topic | Item # | Checklist item | Location where item is reported |
| --- | --- | --- | --- |
| **TITLE** |  |  |  |
| Title | 1 | Identify the report as a systematic review. | 1 |
| **ABSTRACT** |  |  |  |
| Abstract | 2 | See the PRISMA 2020 for Abstracts checklist. | 7-8 |
| **INTRODUCTION** |  |  |  |
| Rationale | 3 | Describe the rationale for the review in the context of existing knowledge. | 9 |
| Objectives | 4 | Provide an explicit statement of the objective(s) or question(s) the review addresses. | 9 |
| **METHODS** |  |  |  |
| Eligibility criteria | 5 | Specify the inclusion and exclusion criteria for the review and how studies were grouped for the syntheses. | 10 |
| Information sources | 6 | Specify all databases, registers, websites, organisations, reference lists and other sources searched or consulted to identify studies. Specify the date when each source was last searched or consulted. | 10 |
| Search strategy | 7 | Present the full search strategies for all databases, registers and websites, including any filters and limits used. | Supp. Methods 1 |
| Selection process | 8 | Specify the methods used to decide whether a study met the inclusion criteria of the review, including how many reviewers screened each record and each report retrieved, whether they worked independently, and if applicable, details of automation tools used in the process. | 10 |
| Data collection process | 9 | Specify the methods used to collect data from reports, including how many reviewers collected data from each report, whether they worked independently, any processes for obtaining or confirming data from study investigators, and if applicable, details of automation tools used in the process. | 10-11 |
| Data items | 10a | List and define all outcomes for which data were sought. Specify whether all results that were compatible with each outcome domain in each study were sought (e.g. for all measures, time points, analyses), and if not, the methods used to decide which results to collect. | 11 |
|  | 10b | List and define all other variables for which data were sought (e.g. participant and intervention characteristics, funding sources). Describe any assumptions made about any missing or unclear information. | 11 |
| Study risk of bias assessment | 11 | Specify the methods used to assess risk of bias in the included studies, including details of the tool(s) used, how many reviewers assessed each study and whether they worked independently, and if applicable, details of automation tools used in the process. | 12 |
| Effect measures | 12 | Specify for each outcome the effect measure(s) (e.g. risk ratio, mean difference) used in the synthesis or presentation of results. |  |
| Synthesis methods | 13a | Describe the processes used to decide which studies were eligible for each synthesis (e.g. tabulating the study intervention characteristics and comparing against the planned groups for each synthesis (item #5)). | 13 |
|  | 13b | Describe any methods required to prepare the data for presentation or synthesis, such as handling of missing summary statistics, or data conversions. | 13-14 |
|  | 13c | Describe any methods used to tabulate or visually display results of individual studies and syntheses. | Table 4 |
|  | 13d | Describe any methods used to synthesize results and provide a rationale for the choice(s). If meta-analysis was performed, describe the model(s), method(s) to identify the presence and extent of statistical heterogeneity, and software package(s) used. | 11 |
|  | 13e | Describe any methods used to explore possible causes of heterogeneity among study results (e.g. subgroup analysis, meta-regression). | 11 |
|  | 13f | Describe any sensitivity analyses conducted to assess robustness of the synthesized results. | 11 |
| Reporting bias assessment | 14 | Describe any methods used to assess risk of bias due to missing results in a synthesis (arising from reporting biases). | 11 |

Supplementary material: PRISMA 2020 Checklist
| Section and Topic | Item # | Checklist item | Location where item is reported |
| --- | --- | --- | --- |
| Certainty assessment | 15 | Describe any methods used to assess certainty (or confidence) in the body of evidence for an outcome. | 11-12 |
| **RESULTS** |  |  |  |
| Study selection | 16a | Describe the results of the search and selection process, from the number of records identified in the search to the number of studies included in the review, ideally using a flow diagram. | 12 |
|  | 16b | Cite studies that might appear to meet the inclusion criteria, but which were excluded, and explain why they were excluded. | Figure 1 |
| Study characteristics | 17 | Cite each included study and present its characteristics. | Table 3 |
| Risk of bias in studies | 18 | Present assessments of risk of bias for each included study. | Table 2 / figure 2 |
| Results of individual studies | 19 | For all outcomes, present, for each study: (a) summary statistics for each group (where appropriate) and (b) an effect estimate and its precision (e.g. confidence/credible interval), ideally using structured tables or plots. | Table 4 / page 14 |
| Results of syntheses | 20a | For each synthesis, briefly summarise the characteristics and risk of bias among contributing studies. | Table 4 / page 14 |
|  | 20b | Present results of all statistical syntheses conducted. If meta-analysis was done, present for each the summary estimate and its precision (e.g. confidence/credible interval) and measures of statistical heterogeneity. If comparing groups, describe the direction of the effect. | Table 4 / page 14 |
|  | 20c | Present results of all investigations of possible causes of heterogeneity among study results. | 14 |
|  | 20d | Present results of all sensitivity analyses conducted to assess the robustness of the synthesized results. | 14 |
| Reporting biases | 21 | Present assessments of risk of bias due to missing results (arising from reporting biases) for each synthesis assessed. | Figure 3 |
| Certainty of evidence | 22 | Present assessments of certainty (or confidence) in the body of evidence for each outcome assessed. | 14 |
| **DISCUSSION** |  |  |  |
| Discussion | 23a | Provide a general interpretation of the results in the context of other evidence. | 14-15 |
|  | 23b | Discuss any limitations of the evidence included in the review. | 15-17 |
|  | 23c | Discuss any limitations of the review processes used. | 16-17 |
|  | 23d | Discuss implications of the results for practice, policy, and future research. | 17-18 |
| **OTHER INFORMATION** |  |  |  |
| Registration and protocol | 24a | Provide registration information for the review, including register name and registration number, or state that the review was not registered. | 9 |
|  | 24b | Indicate where the review protocol can be accessed, or state that a protocol was not prepared. | 9 |
|  | 24c | Describe and explain any amendments to information provided at registration or in the protocol. | n/a |
| Support | 25 | Describe sources of financial or non-financial support for the review, and the role of the funders or sponsors in the review. | 18 |
| Competing interests | 26 | Declare any competing interests of review authors. | COI forms |
| Availability of data, code and other materials | 27 | Report which of the following are publicly available and where they can be found: template data collection forms; data extracted from included studies; data used for all analyses; analytic code; any other materials used in the review. |  |

*From:* Page MJ, McKenzie JE, Bossuyt PM, Boutron I, Hoffmann TC, Mulrow CD, et al. The PRISMA 2020 statement: an updated guideline for reporting systematic reviews. BMJ 2021;372:n71. doi: 10.1136/bmj.n71

## Supplementary methods: full search strategy

### Medline

(Monkeypox OR monkeypox virus OR monkey pox OR MPV) AND ((neurol* OR nervous OR brain OR CNS OR encephal* OR mening* OR myeli* OR myalg* OR ADEM OR ataxi* OR dysphasi* OR aphasi* OR stroke OR guillain-barre OR Miller-Fisher OR paresis OR palsy OR cerebr* OR crani* OR epilep* OR seizure or headache* OR migraine* OR demyeli* OR neuroimag* OR neurotrop* OR neuroinvas* OR neuropath* OR cerebrospinal* or cerebro-spinal OR CSF OR*deliri* OR sleep OR insomnia OR somnolence OR hypersomnolence OR parasomnia OR "movement disorder" OR neuropsych* OR dement* OR cogniti* OR irritability OR hallucinat* OR delusion* OR apath* OR indifference OR agitat* OR euphori* OR elation OR elated OR disinhibit* OR aggressi* OR amnes* OR catatoni* OR personality OR psycho* OR mental OR mood OR affective OR depress* OR anxi* OR "obsessive compulsive" OR OCD OR "panic disorder" OR post-trauma* OR posttrauma* OR PTSD OR neurosis OR neurotic OR bipolar OR mania OR manic OR schizophreni* OR "intelligence quotient" OR IQ OR "mental retardation" OR "intellectual disability" OR "learning disability" OR autis* OR asperger* OR "attention deficit" OR ADHD OR hyperactivity OR hyperkinetic OR suicid* OR emotion* OR appetite OR fatigu* OR tired* OR confus*).ti,ab

OR

(exp Neurology/ or exp Nervous System/ or exp Nervous System Diseases/ or exp Neurologic Manifestations/ or exp Psychiatry/ or exp Mental Processes/ or exp Behavioral Symptoms/ or exp Psychological Phenomena/ or exp DELIRIUM/ OR exp SLEEP/ OR exp WAKEFULNESS/ OR exp SLEEP/ OR exp "DISORDERS OF EXCESSIVE SOMNOLENCE"/ OR exp PARASOMNIAS/ OR exp "PSYCHOMOTOR DISORDERS"/ OR exp DEMENTIA/ OR exp "NEUROCOGNITIVE DISORDERS"/ OR exp HALLUCINATIONS/ OR exp DELUSIONS/ OR exp APATHY/ OR exp "PSYCHOMOTOR AGITATION"/ OR exp EUPHORIA/ OR exp AGGRESSION/ OR exp AMNESIA/ OR exp CATATONIA/ OR exp "PERSONALITY DISORDERS"/ OR exp "SCHIZOPHRENIA SPECTRUM AND OTHER PSYCHOTIC DISORDERS"/ OR exp "MENTAL DISORDERS"/ OR exp "MOOD DISORDERS"/ OR exp DEPRESSION/ OR exp ANXIETY/ OR exp "ANXIETY DISORDERS"/ OR exp "OBSESSIVECOMPULSIVE DISORDER"/ OR exp "PANIC DISORDER"/ OR exp "STRESS DISORDERS, POST-TRAUMATIC"/ OR exp "BIPOLAR AND RELATED DISORDERS"/ OR exp SCHIZOPHRENIA/ OR exp "INTELLECTUAL DISABILITY"/ OR exp "AUTISM SPECTRUM DISORDER"/ OR exp "ASPERGER SYNDROME"/ OR exp "ATTENTION DEFICIT AND DISRUPTIVE BEHAVIOR DISORDERS"/ OR exp "ATTENTION DEFICIT DISORDER WITH HYPERACTIVITY"/ OR exp "MOTOR ACTIVITY"/ OR exp SUICIDE/ OR exp EMOTIONS/ OR exp APPETITE/ OR exp "FEEDING AND EATING DISORDERS"/ OR exp FATIGUE/ OR exp CONFUSION/)) [Humans]

### Embase

(Monkeypox OR monkeypox virus OR monkey pox OR MPV) AND ((neurol* OR nervous OR brain OR CNS OR encephal* OR mening* OR myeli* OR myalg* OR ADEM OR ataxi* OR dysphasi* OR aphasi* OR stroke OR guillain-barre OR Miller-Fisher OR paresis OR palsy OR cerebr* OR crani* OR epilep* OR seizure or headache* OR migraine* OR demyeli* OR neuroimag* OR neurotrop* OR neuroinvas* OR neuropath* OR cerebrospinal* or cerebro-spinal OR CSF OR*deliri* OR sleep OR insomnia OR somnolence OR hypersomnolence OR parasomnia OR "movement disorder" OR neuropsych* OR dement* OR cogniti* OR irritability OR hallucinat* OR delusion* OR apath* OR indifference OR agitat* OR euphori* OR elation OR elated OR disinhibit* OR aggressi* OR amnes* OR catatoni* OR personality OR psycho* OR mental OR mood OR affective OR depress* OR anxi* OR "obsessive compulsive" OR OCD OR "panic disorder" OR post-trauma* OR posttrauma* OR PTSD OR neurosis OR neurotic OR bipolar OR mania OR manic OR schizophreni* OR "intelligence quotient" OR IQ OR "mental retardation" OR "intellectual disability" OR "learning disability" OR autis* OR asperger* OR "attention deficit" OR ADHD OR hyperactivity OR hyperkinetic OR suicid* OR emotion* OR appetite OR fatigu* OR tired* OR confus*).ti,ab

OR

(exp Neuroscience/ or exp Nervous System/ or exp Neurologic Disease/ or exp Psychiatry/ or exp Behavior/ or exp Mental Function/ or exp Psychophysiology/ or exp DELIRIUM/ OR exp "SLEEP DISORDER"/ OR exp INSOMNIA/ OR exp SOMNOLENCE/ OR exp HYPERSOMNIA/ OR exp PARASOMNIA/ OR exp "MOTOR DYSFUNCTION"/ OR exp DEMENTIA/ OR exp "COGNITIVE DEFECT"/ OR exp IRRITABILITY/ OR exp HALLUCINATION/ OR exp DELUSION/ OR exp APATHY/ OR exp AGITATION/ OR exp EUPHORIA/ OR exp AGGRESSION/ OR exp AMNESIA/ OR exp CATATONIA/ OR exp "PERSONALITY DISORDER"/ OR exp PSYCHOSIS/ OR exp "MENTAL DISEASE"/ OR exp MOOD/ OR exp "MOOD DISORDER"/ OR exp DEPRESSION/ OR exp 7 ANXIETY/ OR exp "ANXIETY DISORDER"/ OR exp "OBSESSIVE COMPULSIVE DISORDER"/ OR exp PANIC/ OR exp "POSTTRAUMATIC STRESS DISORDER"/ OR exp NEUROSIS/ OR exp "BIPOLAR DISORDER"/ OR exp MANIA/ OR exp PSYCHOSIS/ OR exp "SCHIZOPHRENIA SPECTRUM DISORDER"/ OR exp SCHIZOPHRENIA/ OR exp "INTELLIGENCE QUOTIENT"/ OR exp "MENTAL DISEASE"/ OR exp "INTELLECTUAL IMPAIRMENT"/ OR exp "DISORDERS OF HIGHER CEREBRAL FUNCTION"/ OR exp "LEARNING DISORDER"/ OR exp AUTISM/ OR exp "ATTENTION DEFICIT DISORDER"/ OR exp HYPERACTIVITY/ OR exp "PSYCHOMOTOR DISORDER"/ OR exp HYPERKINESIA/ OR exp SUICIDE/ OR exp "SUICIDAL BEHAVIOR"/ OR exp "SUICIDE ATTEMPT"/ OR exp EMOTION/ OR exp APPETITE/ OR exp "APPETITE DISORDER"/ OR exp FATIGUE/ OR exp CONFUSION))

[Humans]

### Psychinfo

(Monkeypox OR monkeypox virus OR monkey pox OR MPV)

AND

((neurol* OR nervous OR brain OR CNS OR encephal* OR mening* OR myeli* OR myalg* OR ADEM OR ataxi* OR dysphasi* OR aphasi* OR stroke OR guillain-barre OR Miller-Fisher OR paresis OR palsy OR cerebr* OR crani* OR epilep* OR seizure or headache* OR migraine* OR demyeli* OR neuroimag* OR neurotrop* OR neuroinvas* OR neuropath* OR cerebrospinal* or cerebro-spinal OR CSF OR*deliri* OR sleep OR insomnia OR somnolence OR hypersomnolence OR parasomnia OR "movement disorder" OR neuropsych* OR dement* OR cogniti* OR irritability OR hallucinat* OR delusion* OR apath* OR indifference OR agitat* OR euphori* OR elation OR elated OR disinhibit* OR aggressi* OR amnes* OR catatoni* OR personality OR psycho* OR mental OR mood OR affective OR depress* OR anxi* OR "obsessive compulsive" OR OCD OR "panic disorder" OR post-trauma* OR posttrauma* OR PTSD OR neurosis OR neurotic OR bipolar OR mania OR manic OR schizophreni* OR "intelligence quotient" OR IQ OR "mental retardation" OR "intellectual disability" OR "learning disability" OR autis* OR asperger* OR "attention deficit" OR ADHD OR hyperactivity OR hyperkinetic OR suicid* OR emotion* OR appetite OR fatigu* OR tired* OR confus*).ti,ab OR (exp Psychiatry/ OR exp Sensory System Disorders/ OR exp Sense Organ Disorders/ OR exp Nervous System Disorders/ OR exp Neurosciences/ or exp Emotional States/ OR exp DELIRIUM/ OR exp "NEUROCOGNITIVE DISORDERS"/ OR exp "SLEEP WAKE DISORDERS"/ OR exp INSOMNIA/ OR exp HYPERSOMNIA/ OR exp "MOVEMENT DISORDERS"/ OR exp DEMENTIA/ OR exp "COGNITIVE ABILITY"/ OR exp "COGNITIVE IMPAIRMENT"/ OR exp "NEUROCOGNITIVE DISORDERS"/ OR exp IRRITABILITY/ OR exp HALLUCINATIONS/ OR exp DELUSIONS/ OR exp APATHY/ OR exp AGITATION/ OR exp EUPHORIA/ OR exp "BEHAVIORAL DISINHIBITION"/ OR exp AMNESIA/ OR exp CATATONIA/ OR exp PERSONALITY/ OR exp "PERSONALITY DISORDERS"/ OR exp PSYCHOSIS/ OR exp "MENTAL DISORDERS"/ OR exp EMOTIONS/ OR exp "AFFECTIVE DISORDERS"/ OR exp "DEPRESSION (EMOTION)"/ OR exp ANXIETY/ OR exp "ANXIETY DISORDERS"/ OR exp "OBSESSIVE COMPULSIVE DISORDER"/ OR exp "PANIC DISORDER"/ OR exp "POSTTRAUMATIC STRESS"/ OR exp NEUROSIS/ OR exp "BIPOLAR DISORDER"/ OR exp MANIA/ OR exp SCHIZOPHRENIA/ OR exp "INTELLIGENCE QUOTIENT"/ OR exp "NEURODEVELOPMENTAL DISORDERS"/ OR exp "INTELLECTUAL DEVELOPMENT DISORDER"/ OR exp "AUTISM SPECTRUM DISORDERS"/ OR exp "ATTENTION DEFICIT DISORDER"/ OR exp HYPERKINESIS/ OR exp SUICIDE/ OR exp EMOTIONS/ OR exp APPETITE/ OR exp "EATING DISORDERS"/ OR exp FATIGUE/ OR exp "MENTAL CONFUSION"/)

[Population Human]

### MedRxiv

(Monkeypox OR monkeypox OR monkey pox OR MPV)

In categories psychiatry and clinical psychology; neurology

